# Integrating Real-Time Location Systems with Electronic Medical Records: A Machine Learning Approach for In-Hospital Fall Risk Prediction

**DOI:** 10.1101/2024.03.11.24304095

**Authors:** Dong Won Kim, Jihoon Seo, Sujin Kwon, Chan Min Park, Changho Han, Yujeong Kim, Dukyong Yoon, Kyoung Min Kim

## Abstract

Hospital falls are the most prevalent adverse event in healthcare, posing significant risks to patient health outcomes and institutional care quality. The effectiveness of several fall prediction models currently in use is limited by various clinical factors. This study explored the efficacy of merging real-time location system (RTLS) data with clinical information to enhance the accuracy of in-hospital fall predictions. The model performances were compared based on the clinical data, RTLS data, and a hybrid approach using various evaluation metrics. The RTLS and integrated clinical data were obtained from 22,201 patients between March 2020 and June 2022. From the initial cohort, 118 patients with falls and 443 patients without falls were included. Predictive models were developed using the XGBoost algorithm across three distinct frameworks: clinical model, RTLS model, and clinical + RTLS model. The model performance was evaluated using metrics, such as AUROC, AUPRC, accuracy, PPV, sensitivity, specificity, and F1 score. Shapley additive explanation values were used to enhance the model interpretability. The clinical model yielded an AUROC of 0.813 and AUPRC of 0.407. The RTLS model demonstrated superior fall prediction capabilities, with an AUROC of 0.842 and AUPRC of 0.480. The clinical + RTLS model excelled further, achieving an AUROC of 0.853 and AUPRC of 0.497. Feature importance analysis revealed that movement patterns of patients on the last day of their stay were significantly associated with falls, together with elevated RDW levels, sedative administration, age. This study underscored the advantages of combining RTLS data with clinical information to predict in-hospital falls more accurately. This innovative technology-driven approach may enhance early fall risk detection during hospitalization, potentially preventing falls, improving patient safety, and contributing to more efficient healthcare delivery.

## 1. Introduction

Managing patient safety and hospital operations is crucial in the healthcare domain. In-hospital falls are among the most common incidents that cause concern in healthcare facilities. This issue not only affects patient health outcomes but also has substantial economic implications. For instance, in the United States, the overall medical expenditure for fatal falls is estimated to be $754 million^1^. This figure underscores the severe impact of falls on healthcare systems worldwide. Reportedly, 15–50% patients suffer fall-related injuries, out of which 1–10% are severe, including fractures^2–5^. This scenario is prevalent in other countries, where 25–55% of the nursing home patients experience falls, leading to significant injuries such as fractures, brain hemorrhage or even death, as seen in 1.2–16.2% of cases in South Korea being fatal^6^. These statistics highlight that falls are not mere accidents, but significant global risk factors causing severe secondary accidents.

To minimize falls, several tools have been used to assess their risks upon admission. However, these methods are constrained by their dependence on subjective assessments and patients’ recall^7–10^. Furthermore, these tools are limited in their ability to assess the changes of patient’s physical ability, which is directly related to fall probability. The hospitalized patients may experience significant alterations in their biological conditions and often spend most of their time in beds following admission, leading to decreased physical activity and reduced gait speed, which directly increases the risk of falls^11–14^. However, a notable limitation of conventional tools is their ineffectiveness in dynamically reflecting changes in patient conditions. Therefore, novel objective tools capable of continuous monitoring and adaptation to the evolving physical abilities and conditions of patients are required^15,16^.

Among the various indicators of a patient’s physical abilities, elements such as the amount of movement, gait speed, and degree of active movement are directly related to falls^13,14^. A real-time location system (RTLS) is a technology that detects and records the location of sensors in real time and has various applications in medical and healthcare settings, including infection path tracking^17^. Its ability to continuously collect time-based location data makes it suitable for objectively tracking changes in patient movements and physical activity. Additionally, electronic medical records (EMRs) store a wealth of objective information on medical conditions that may reflect a patient’s vulnerability. Considering the traceability of RTLS for physical ability and objective information about clinical situations contained in EMR, this study aims to overcome the aforementioned issues by developing a fall-prediction machine-learning model. This integrated approach develops a comprehensive and innovative fall-prediction tool that offers an advanced method for enhancing patient safety and healthcare management.

## 2. Results

### 2.1 Study design

Fig. 1 shows the framework of this study, which spanned from March 1, 2020 to June 30, 2022 and developed a predictive model for falls during hospital admissions using the EMR and RTLS data from the Yongin Severance Hospital. These data were obtained from patients who satisfied the inclusion criteria (Fig. 1a). The EMR data included 27 standard features procured during the initial phase of patient admission (Fig. 1b). The RTLS data, encompassing geographical coordinates and timestamps, were used to generate 28 additional features per patient, reflecting their physical activities (Fig. 1c). This comprehensive approach resulted in a dataset with 55 features, which served as the foundation for machine learning analysis. The Extreme Gradient Boost (XGBoost) algorithm was applied based on a 10-fold cross-validation method for fall prediction during hospital stay (Fig. 1d).

**Fig. 1.**
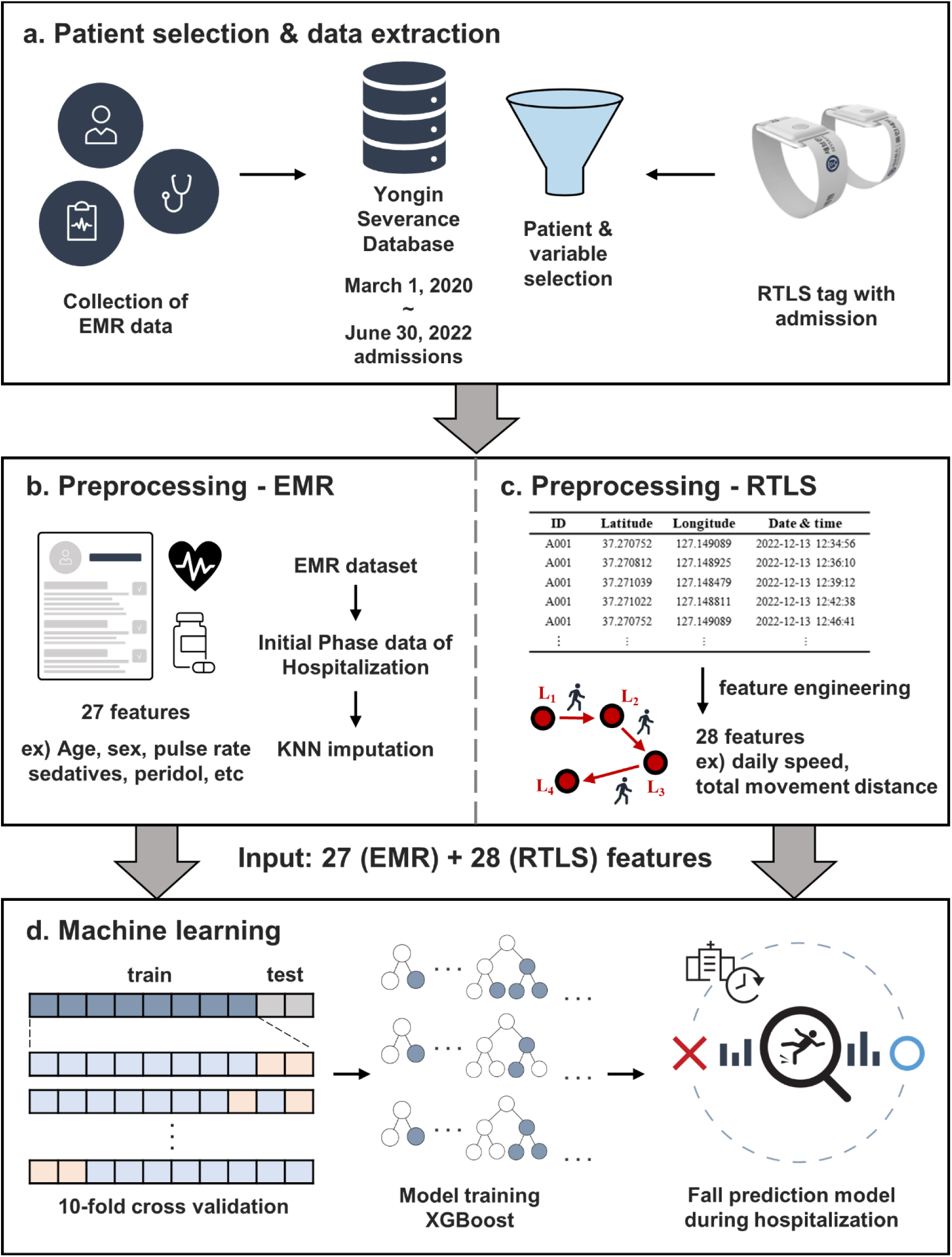
Overall research flow. a, Patient selection and data extraction. Selection of study participants admitted to Yongin Severance Hospital and extraction of data from the EMR and RTLS databases. b, Preprocessing 27 features from the EMR database. c, Preprocessing 28 features from the RTLS database reflecting the physical activities of patients. d, Development of a machine-learning-based prediction model for falls during hospitalization based on the preprocessed data by employing the XGBoost algorithm. EMR: electronic medical record, RTLS: real-time location system, KNN imputation: k-nearest neighbor imputation, XGBoost: the extreme gradient boost algorithm.

### 2.2 Differences in baseline clinical characteristics and RTLS profiles based on the fall experience

The study cohort comprised 118 patients with falls and 443 patients without falls (Fig. 2). Statistical comparison of baseline characteristics between fall and non-fall groups revealed significant differences in several clinical and RTLS features (Table 1 and Supplementary Table 1). In Table 1, patients experiencing falls were predominantly older males with longer hospital stays and higher dosage of sedatives or peridol. They were primarily admitted for medical treatment and exhibited higher rates of intensive care unit (ICU) admission. Regarding biochemical parameters, there were no significant differences between the groups for most variables; however, the red cell distribution width (RDW) values were notably higher in the fall group than in the non-fall group. The RTLS data revealed significant disparities in the daily speed on the last day, as well as in the mean and median daily speed, indicating a slower daily speed in the fall group than in the non-fall group during hospital stay (Supplementary Table 1). Here, the terms "Daily top 20% movement ratio" and "Daily top 50% movement ratio" refer to the proportion of time during a day when a patient’s velocity is within the highest 20% and 50%, respectively, among all patient velocity measurements. It was found that for the daily top 20% movement ratio, there were notable differences not only in the average and median values throughout their hospital stay but also in the values recorded on the last day between the groups, indicating lower proportion of daily top 20% movement ratio in fall groups. Similarly, significant differences were also observed for the daily top 50% movement on the last day and in its average value between the groups (Supplementary Table 1). These underscores a marked reduction in overall physical activity levels in the fall group compared to the non-fall group throughout their hospital stay.

**Fig. 2.**
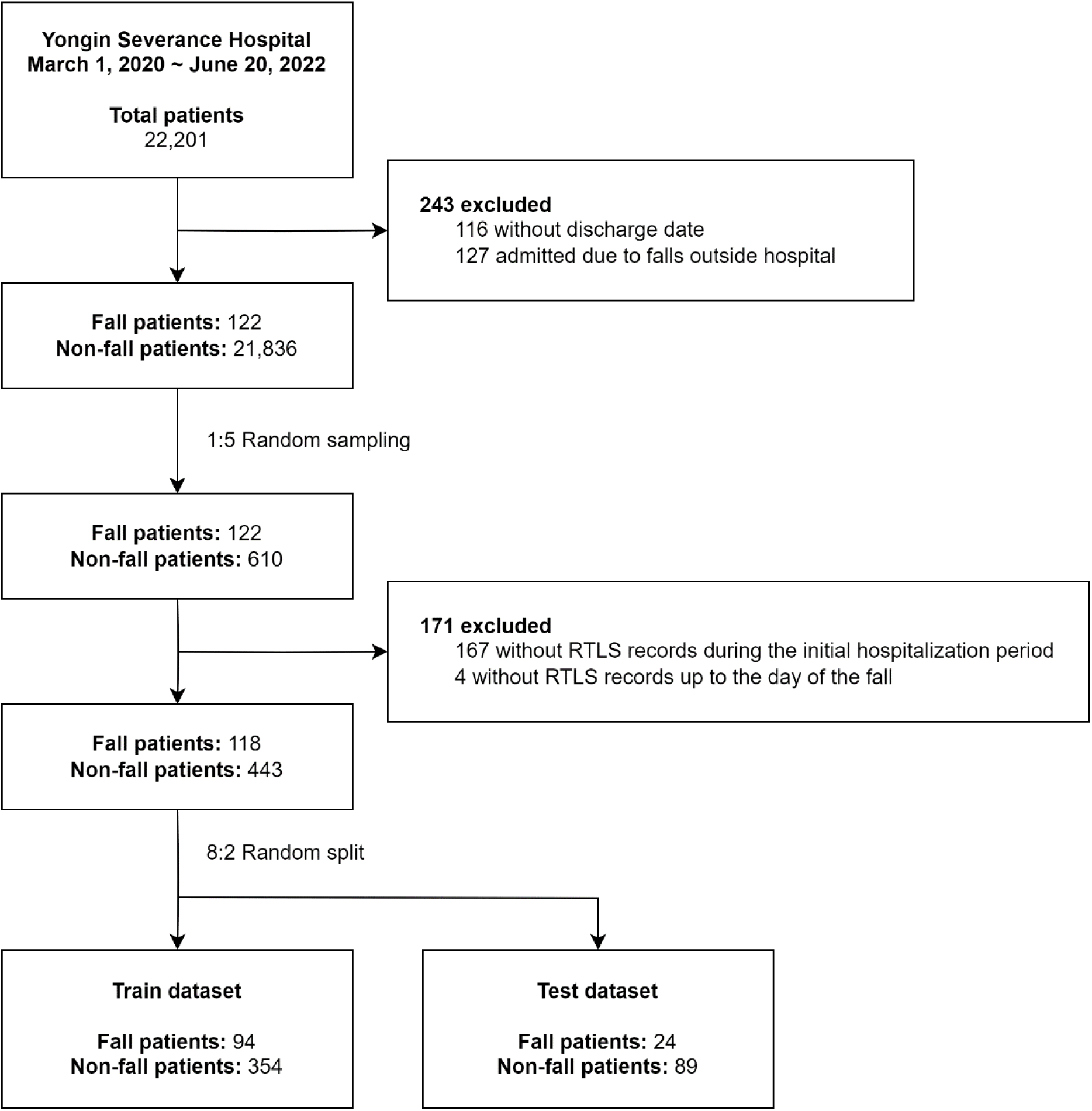
Patient selection flowchart. RTLS: real-time location system.

**Table 1.**
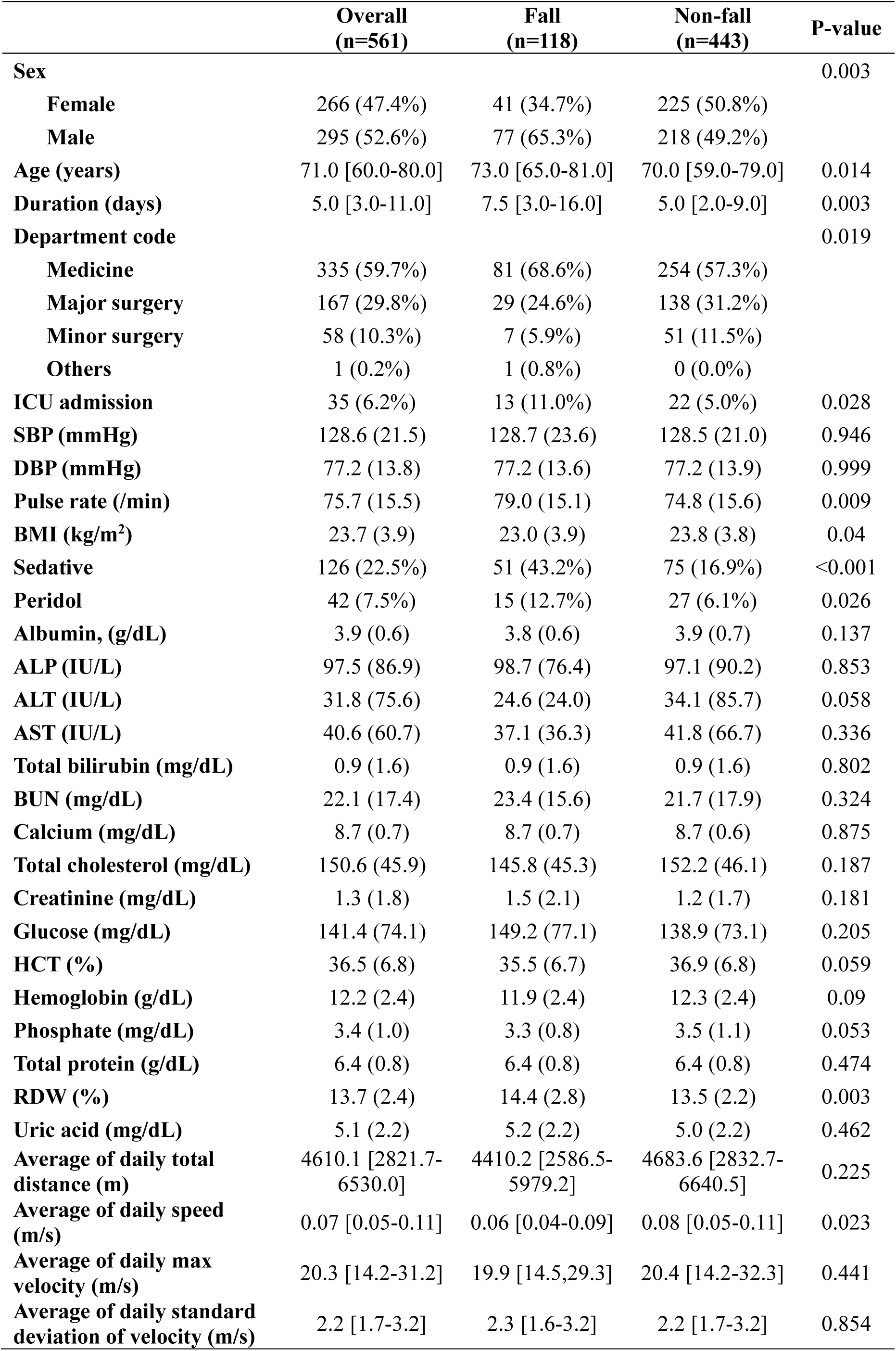
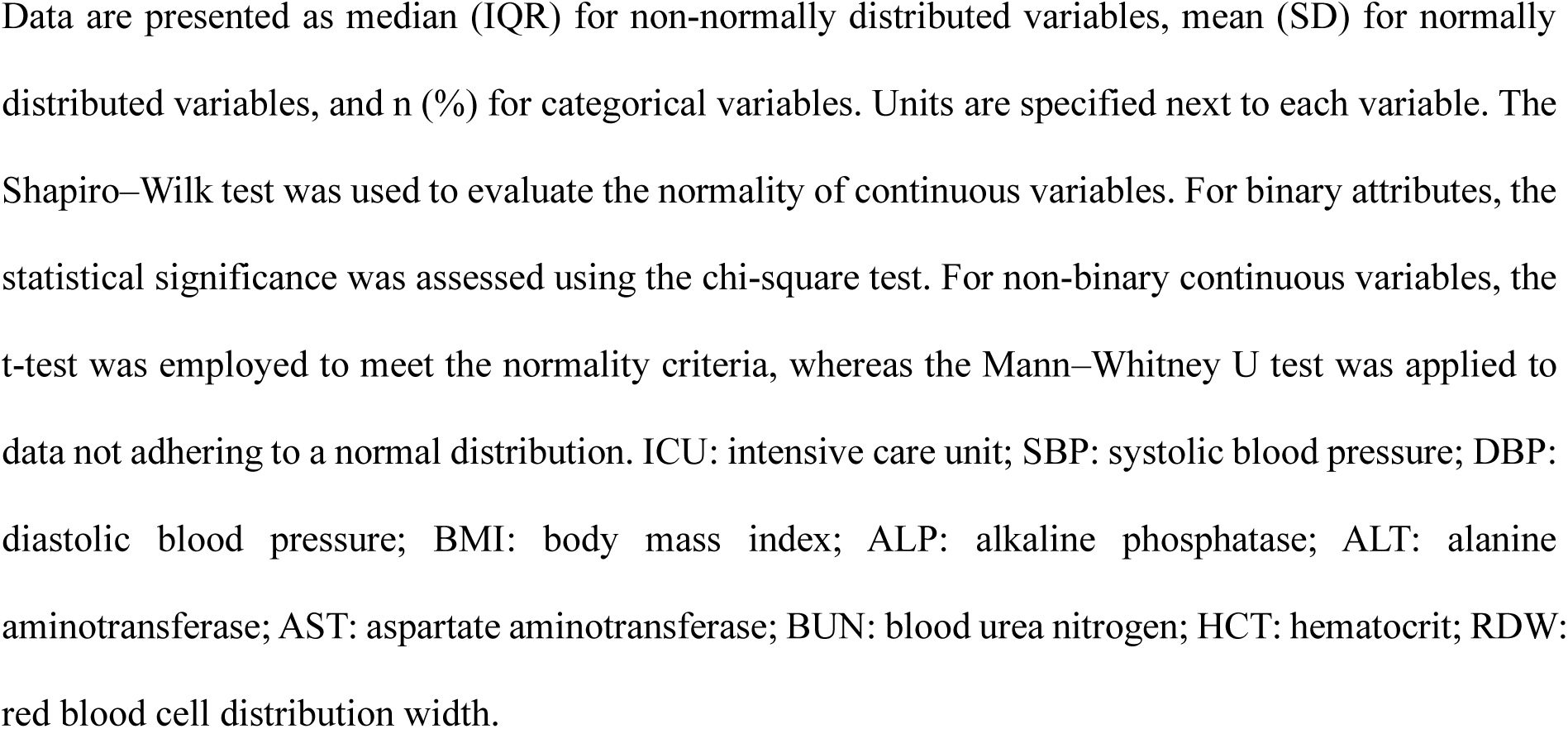
Baseline characteristics of patients.

|  | **Overall**<br>**(n=561)** | **Fall**<br>**(n=118)** | **Non-fall**<br>**(n=443)** | **P-value** |
| --- | --- | --- | --- | --- |
| **Sex** |  |  |  | 0.003 |
| **Female** | 266 (47.4%) | 41 (34.7%) | 225 (50.8%) |  |
| **Male** | 295 (52.6%) | 77 (65.3%) | 218 (49.2%) |  |
| **Age (years)** | 71.0 [60.0-80.0] | 73.0 [65.0-81.0] | 70.0 [59.0-79.0] | 0.014 |
| **Duration (days)** | 5.0 [3.0-11.0] | 7.5 [3.0-16.0] | 5.0 [2.0-9.0] | 0.003 |
| **Department code** |  |  |  | 0.019 |
| **Medicine** | 335 (59.7%) | 81 (68.6%) | 254 (57.3%) |  |
| **Major surgery** | 167 (29.8%) | 29 (24.6%) | 138 (31.2%) |  |
| **Minor surgery** | 58 (10.3%) | 7 (5.9%) | 51 (11.5%) |  |
| **Others** | 1 (0.2%) | 1 (0.8%) | 0 (0.0%) |  |
| **ICU admission** | 35 (6.2%) | 13 (11.0%) | 22 (5.0%) | 0.028 |
| **SBP (mmHg)** | 128.6 (21.5) | 128.7 (23.6) | 128.5 (21.0) | 0.946 |
| **DBP (mmHg)** | 77.2 (13.8) | 77.2 (13.6) | 77.2 (13.9) | 0.999 |
| **Pulse rate (/min)** | 75.7 (15.5) | 79.0 (15.1) | 74.8 (15.6) | 0.009 |
| **BMI (kg/m<sup>2</sup>)** | 23.7 (3.9) | 23.0 (3.9) | 23.8 (3.8) | 0.04 |
| **Sedative** | 126 (22.5%) | 51 (43.2%) | 75 (16.9%) | <0.001 |
| **Peridol** | 42 (7.5%) | 15 (12.7%) | 27 (6.1%) | 0.026 |
| **Albumin, (g/dL)** | 3.9 (0.6) | 3.8 (0.6) | 3.9 (0.7) | 0.137 |
| **ALP (IU/L)** | 97.5 (86.9) | 98.7 (76.4) | 97.1 (90.2) | 0.853 |
| **ALT (IU/L)** | 31.8 (75.6) | 24.6 (24.0) | 34.1 (85.7) | 0.058 |
| **AST (IU/L)** | 40.6 (60.7) | 37.1 (36.3) | 41.8 (66.7) | 0.336 |
| **Total bilirubin (mg/dL)** | 0.9 (1.6) | 0.9 (1.6) | 0.9 (1.6) | 0.802 |
| **BUN (mg/dL)** | 22.1 (17.4) | 23.4 (15.6) | 21.7 (17.9) | 0.324 |
| **Calcium (mg/dL)** | 8.7 (0.7) | 8.7 (0.7) | 8.7 (0.6) | 0.875 |
| **Total cholesterol (mg/dL)** | 150.6 (45.9) | 145.8 (45.3) | 152.2 (46.1) | 0.187 |
| **Creatinine (mg/dL)** | 1.3 (1.8) | 1.5 (2.1) | 1.2 (1.7) | 0.181 |
| **Glucose (mg/dL)** | 141.4 (74.1) | 149.2 (77.1) | 138.9 (73.1) | 0.205 |
| **HCT (%)** | 36.5 (6.8) | 35.5 (6.7) | 36.9 (6.8) | 0.059 |
| **Hemoglobin (g/dL)** | 12.2 (2.4) | 11.9 (2.4) | 12.3 (2.4) | 0.09 |
| **Phosphate (mg/dL)** | 3.4 (1.0) | 3.3 (0.8) | 3.5 (1.1) | 0.053 |
| **Total protein (g/dL)** | 6.4 (0.8) | 6.4 (0.8) | 6.4 (0.8) | 0.474 |
| **RDW (%)** | 13.7 (2.4) | 14.4 (2.8) | 13.5 (2.2) | 0.003 |
| **Uric acid (mg/dL)** | 5.1 (2.2) | 5.2 (2.2) | 5.0 (2.2) | 0.462 |
| **Average of daily total distance (m)** | 4610.1 [2821.7-6530.0] | 4410.2 [2586.5-5979.2] | 4683.6 [2832.7-6640.5] | 0.225 |
| **Average of daily speed (m/s)** | 0.07 [0.05-0.11] | 0.06 [0.04-0.09] | 0.08 [0.05-0.11] | 0.023 |
| **Average of daily max velocity (m/s)** | 20.3 [14.2-31.2] | 19.9 [14.5,29.3] | 20.4 [14.2-32.3] | 0.441 |
| **Average of daily standard deviation of velocity (m/s)** | 2.2 [1.7-3.2] | 2.3 [1.6-3.2] | 2.2 [1.7-3.2] | 0.854 |

Data are presented as median (IQR) for non-normally distributed variables, mean (SD) for normally distributed variables, and n (%) for categorical variables. Units are specified next to each variable. The Shapiro–Wilk test was used to evaluate the normality of continuous variables. For binary attributes, the statistical significance was assessed using the chi-square test. For non-binary continuous variables, the t-test was employed to meet the normality criteria, whereas the Mann–Whitney U test was applied to data not adhering to a normal distribution. ICU: intensive care unit; SBP: systolic blood pressure; DBP: diastolic blood pressure; BMI: body mass index; ALP: alkaline phosphatase; ALT: alanine aminotransferase; AST: aspartate aminotransferase; BUN: blood urea nitrogen; HCT: hematocrit; RDW: red blood cell distribution width.

### 2.3 Performance comparison of the three models for fall prediction

The performances of the three models derived from the patient cohorts were compared using the test set. Based on the comparison of average values, the clinical, RTLS, and clinical + RTLS models exhibited area under the receiver operating characteristic curve (AUROC) values of 0.699, 0.813, and 0.847, respectively, indicating statistically significant performance improvements by integrating comprehensive variables. Similarly, the clinical, RTLS, and clinical + RTLS models achieved area under the precision-recall curve (AUPRC) values of 0.423, 0.565, and 0.667, respectively, demonstrating parallel significant differences. The corresponding Brier scores for the models were 0.159, 0.133, and 0.120, with clinical + RTLS model exhibiting the best performance (Fig. 3 and Table 2).

**Fig. 3.**
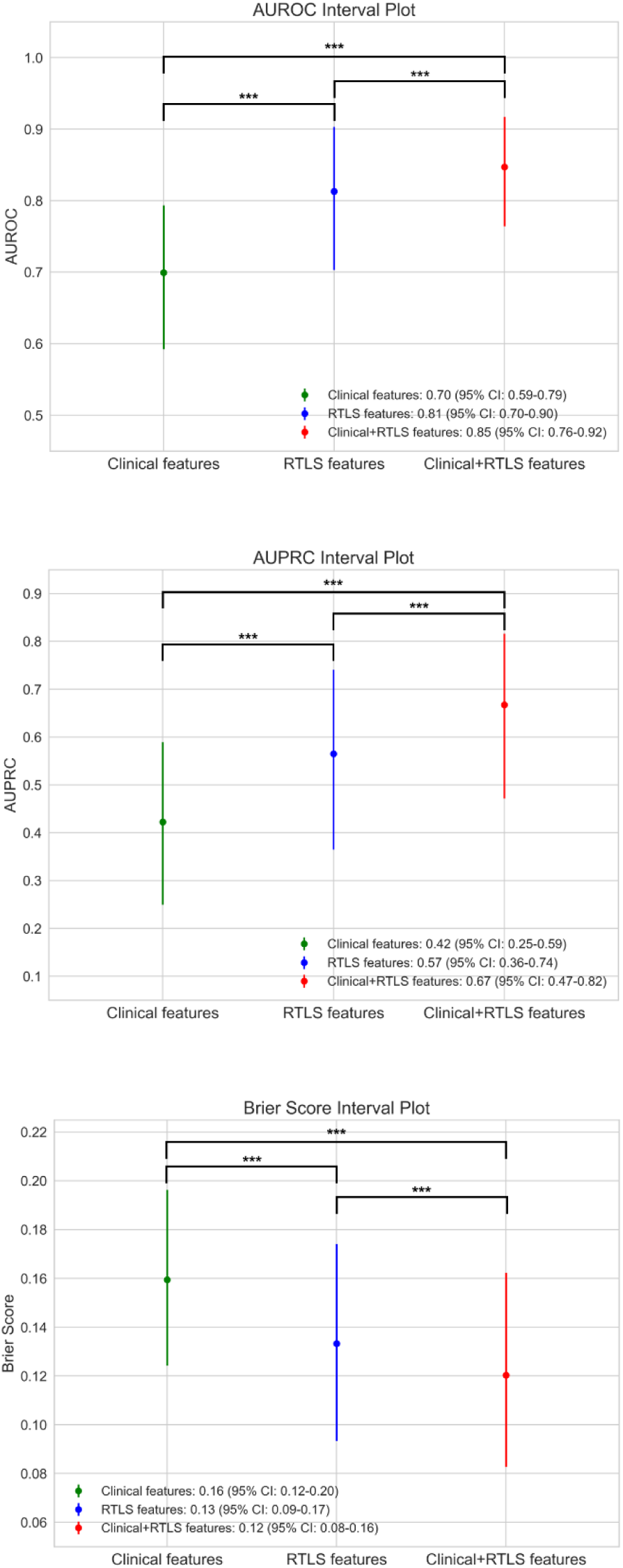
Graphical performance evaluation of the three models. Performance comparison of the three models using AUROC, AUPRC, and Brier score metrics based on 1000 bootstrap resampling. Results are presented with 95% confidence intervals. Statistical significance was assessed using the Kruskal–Wallis test followed by Dunn’s post-hoc test, with significance levels indicated on the graph. The clinical, RTLS, and clinical + RTLS models are denoted in green, blue, and red, respectively. AUROC: area under the receiver operating characteristics curve; AUPRC: area under the precision recall curve; CI: confidence interval.

**Table 2.** Comparative performances of the three models.

| **Model** | **Youden index** | **AUROC** | **AUPRC** | **Brier score** |
| --- | --- | --- | --- | --- |
| Clinical model | 0.298 | 0.699<br>(0.592-0.794) | 0.423<br>(0.250-0.590) | 0.159<br>(0.124-0.196) |
| RTLS model | 0.428 | 0.813<br>(0.703-0.903) | 0.565<br>(0.365-0.741) | 0.133<br>(0.093-0.174) |
| Clinical + RTLS model | 0.376 | 0.847<br>(0.764-0.917) | 0.667<br>(0.472-0.816) | 0.120<br>(0.083-0.162) |

Displayed values are the performance metrics for the clinical, RTLS, and clinical + RTLS models, with 95% confidence intervals (CIs) in parentheses. RTLS: real-time location system; AUROC: area under the receiver operating characteristic curve; AUPRC: area under the precision-recall curve.

Based on other metrics for comparing model performances, the clinical + RTLS model outperformed the other models, exhibiting statistically significant variations and achieving an accuracy of 0.848, positive predictive value (PPV) of 0.681, sensitivity of 0.540, specificity of 0.931, and F1 score of 0.600 (Supplementary Fig. 1 and Supplementary Table 2).

### 2.4 Comparison of the model feature importances between the three models

Fig. 4 shows the feature importance of the clinical + RTLS model, which exhibited the best performance. The significance of each feature was assessed and compared across the remaining models (Supplementary Fig. 2). The predominant features influencing the clinical model (Supplementary Fig. 2a) and the RTLS feature model (Supplementary Fig. 2b) were largely retained in the clinical + RTLS model.

**Fig. 4.**
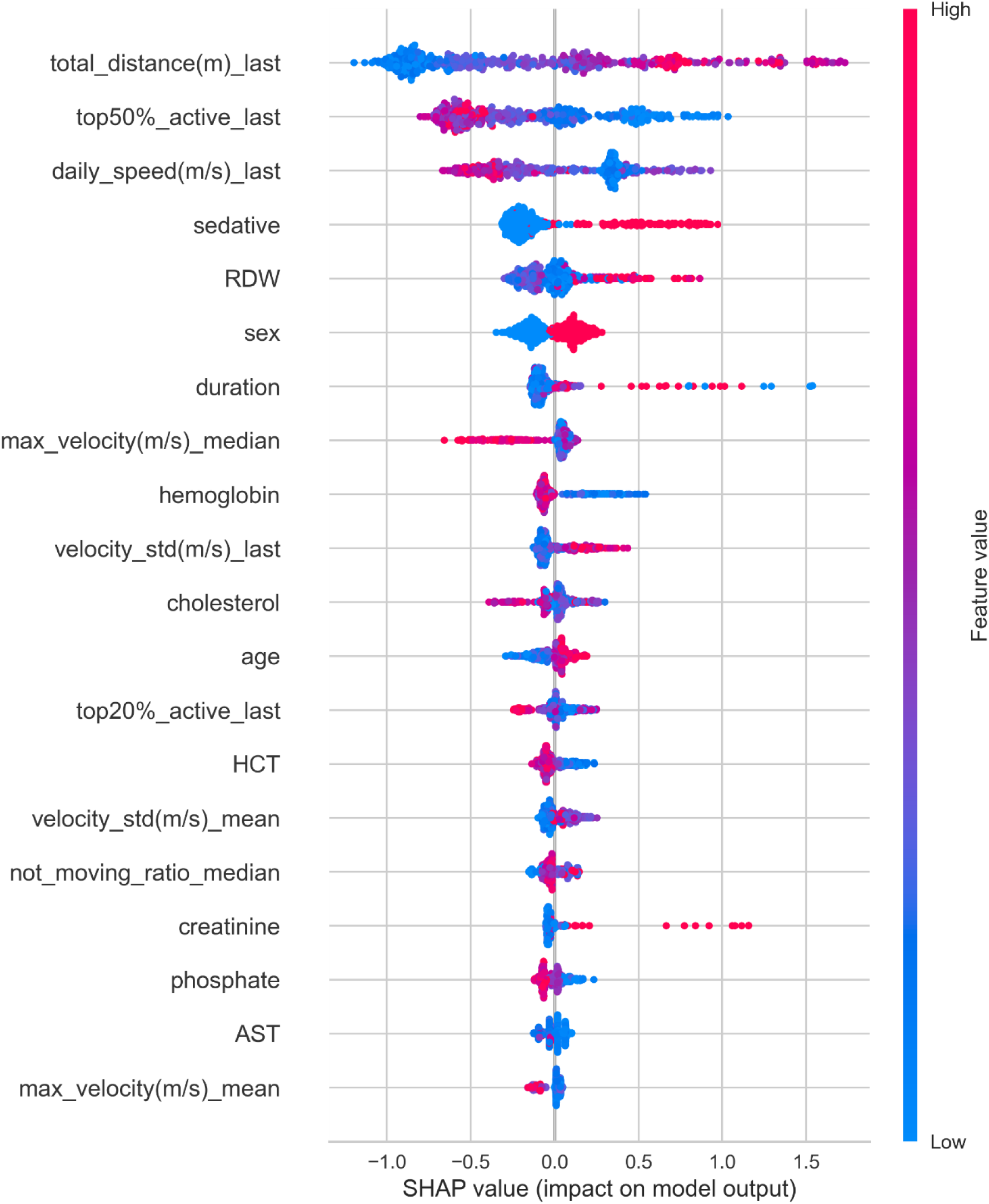
Visualizing feature importance for the combined clinical + RTLS model using SHAP values. Impact of the top 20 features on model prediction, ranked based on SHAP values. The y-axis displays features in the order of decreasing impact, whereas the x-axis quantifies their SHAP values, with the color intensity ranging from blue for lower values to red for higher values. total_distance(m)_last: total distance moved (in meter) on the last day of RTLS recording; top50%_active_last: ratio of time spent in the top 50% of active movements on the last day of RTLS recording; max_velocity(m/s)_median: median of daily maximum velocities recorded throughout hospitalization; velocity_std(m/s)_last: standard deviation of velocities on the last day of RTLS recording; top20%_active_last: ratio of time spent in the top 20% of active movements on the last day of RTLS recording; velocity_std(m/s)_mean: mean of daily velocity standard deviations throughout hospitalization; not_moving_ratio_median: median of the daily ratios of time spent without movement throughout hospitalization; max_velocity(m/s)_mean: mean of daily maximum velocities recorded throughout hospitalization; AST: aspartate aminotransferase; HCT: hematocrit, RDW: red cell distribution width; SHAP value: Shapley additive explanations value.

In the clinical + RTLS model, SHAP analysis identified the daily total distance moved (total_distance), the daily top 50% active movement ratio (top50%_active), and the daily movement speed (daily_speed) as the top three ranked features, with their values on the last day of hospitalization being particularly impactful (Fig. 4). It was found that a greater total distance moved on the last day significantly increased the likelihood of a fall occurring. Conversely, a lower daily top 50% active movement ratio and a slower daily movement speed on the last day were found to be strongly predictive of falls. Here, the daily total distance represents the cumulative distance a patient covers throughout a day, and the daily top 50% active movement ratio quantifies the proportion of time a patient spends in more vigorous activity. A greater total distance moved on the last day significantly increased the likelihood of a fall occurring. Conversely, a lower daily top 50% active movement ratio and a slower daily movement speed on the last day were found to be strongly predictive of falls. Furthermore, the fourth- and fifth-ranked features revealed that the elevated RDW levels of patients who were administered sedative drugs greatly affected the fall incidents.

### 2.5 Comparative evaluation of the model clinical efficacy using net benefit analysis

A decision curve analysis (DCA) comparing the three models revealed that the clinical model, which utilized only EMR variables, exhibited the lowest net benefit and offered minimal clinical advantages (Fig. 5). Contrastingly, the clinical + RTLS model, consistent with its performance in other metrics, exhibited a superior net benefit across various thresholds compared with the other two models.

**Fig. 5.**
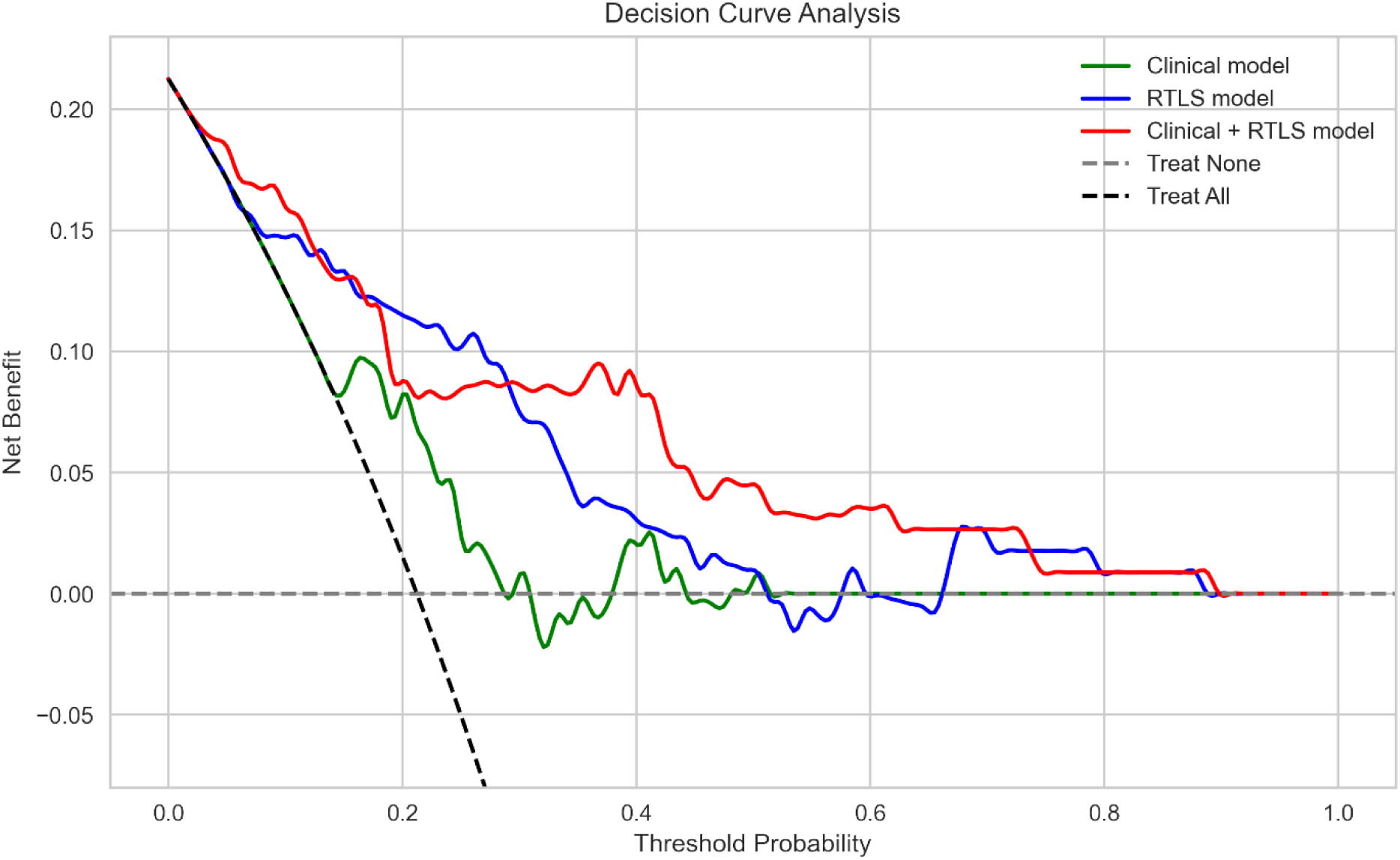
DCA of each model performance. Decision curve analysis comparing the models based on net benefit across decision thresholds. Net benefit quantifies the trade-off between the benefit of true positives and harm of false positives. The clinical, RTLS, and clinical + RTLS models are denoted in green, blue, and red, respectively. RTLS: real-time location system.

## 3. Discussion

The results of this study indicate a significant advancement in the field of fall prediction during hospitalization by utilizing a novel approach that integrates the RTLS variables capturing patient movement with the clinical variables obtained from EMR. This comprehensive model, which considers both sets of variables, outperforms the models using either dataset by itself. The clinical + RTLS model achieved an AUROC of 0.847, AUPRC of 0.667, and Brier score of 0.120, and the clinical benefits of the proposed model are further confirmed by DCA. These findings may significantly enhance fall prevention by facilitating a more convenient and precise prediction of such incidents.

The proposed approach remarkably overcomes the limitations of conventional methods that lack the ability to continuously and objectively measure patient movement^7–9,18,19^. By utilizing real-time data from RTLS, we engineered variables capturing variations in patient movement and incorporated initial clinical scores and medication histories from EMRs to enhance the fall prediction performance. Instead of relying on a single model, this study simultaneously presents three independently developed models based on clinical features, RTLS data, and a combination of these two. Each dataset facilitates commendable predictive performance individually; however, the integrated model demonstrates statistically significant superiority, indicating that the initial clinical measurements and RTLS-derived features contribute to prediction accuracy.

SHAP analysis provides insights into the most influential features of the high-performing clinical + RTLS model. Among the top five features, three were derived from RTLS (daily total distance, daily speed, and daily top 50% active movement ratio) and remaining two were derived from the EMR data, indicating a balanced contribution from the two data sources. The RTLS variables, particularly the daily total distance, represent the total amount of patient movement in a day. Contrastingly, the daily speed represents an average daily measure of the gait speed, a well-known factor associated with falls that is emphasized in conventional methods^13,20–22^. The daily top 50% active movement ratio, which indicates the proportion of time spent on moving at a faster pace, serves as an indirect measure of physical activity, another key factor in determining fall risk^11,23^. These RTLS variables are analyzed across various time points, including initial hospitalization and the day of the fall, revealing that the values on the last day of stay are particularly important. Therefore, the patient movement patterns closer to the time of fall are more critical than those at the beginning of hospitalization. This aligns with the existing research advocating the continuous monitoring of patients to prevent falls^15,16^. Hence, a decrease in physical activity and gait speed on days closer to the fall, particularly in patients with more movement, is correlated with an increased risk of falling.

In addition to RTLS variables, high RDW and sedative use are associated with an increased risk of falls, corroborating the findings from existing research. Sedatives, which are central nervous system depressants typically prescribed for anxiety and sleep disorders, are associated with falls owing to their effects such as slowing reaction times and causing dizziness^24,25^. The relationship between sedative use and fall risk highlights the need for careful management of these medications in hospitalized patients, particularly in those at a higher risk of falls. An increase in this metric may indicate the biological aging process impairing neuromuscular function, which can increase the fall risk as it potentially affects the balance, coordination, and overall physical strength^26^. The correlation between higher RDW and fall risk indicates that this hematological parameter could serve as a useful biomarker for fall risk assessment. RDW has been known to be closely related to aging phenotypes in diverse clinical situations^26–28^. Integrating RDW with the proposed model, along with other clinical and RTLS-derived variables, provides a more comprehensive understanding of the multifactorial nature of fall risk in hospital settings.

The implications of this study can be understood in three ways. First, the model generalizability is emphasized because the study was based on a randomly sampled population of all hospitalized patients in a tertiary care hospital, rather than being limited to specific wards or disease conditions. Second, by capturing the real-time dynamics of patient movements using RTLS, the proposed model overcomes the limitations of conventional fall prediction methods, offering a more accurate and timely assessment of fall risk. Third, the alignment of the key variables of the model, identified via SHAP analysis, with the reported fall risk factors underscores the strong explainability of the model. This congruence validates the fundamental requirement for clinically verified knowledge, thus enhancing the credibility and utility of the model in clinical settings.

The primary limitation of this study is the requirement for multi-cohort validation. The proposed approach, involving 1000 iterations with duplication in the test set, serves as a provisional measure to statistically validate the model performance; however, future research should perform validation across diverse cohorts. Another limitation is data imbalance owing to the varying patient event distributions. Despite mitigating this issue via random sampling, disparities in patient data persisted, which may have affected the accuracy and precision of the findings. The reliance on RTLS and EMR data, while informative, could omit potential environmental or situational factors influencing the fall risk. Further research is necessary to capture these broader aspects and enhance the understanding of fall risk in hospital settings.

To the best of our knowledge, this study is the first to apply RTLS to fall prediction. Advancements in IT technologies have enabled the tracking of patient locations within hospitals using RTLS. This study seeks to overcome the limitations of previous research on falls by leveraging technological progress. By utilizing real-time data on patient location changes, our findings suggest that integrating RTLS with EMR can significantly improve fall-prediction accuracy. This IT-enhanced approach may facilitate the early detection of fall risks during hospitalization, thereby preventing fall incidents, augmenting patient safety, and alleviating the workload of healthcare professionals by implementing automated solutions.

## 4. Methods

### 4.1 Study participants and data source

We conducted a retrospective study by extracting the EMR and RTLS data from 22,201 patients admitted to Yongin Severance Hospital between March 1, 2020, and June 30, 2022. The patients were provided with RTLS-equipped wristbands (Supplementary Fig. 3). The RTLS was designed to update its database only when there was a change in patient’s location, capturing the date, time, latitude, and longitude.

The initial cohort of patients was refined by excluding individuals with unavailable discharge records (n=116) and those admitted because of falls from external sources (n=127), thereby focusing our analysis on in-hospital falls. The patient cohort was divided into the following two groups: those who experienced falls (fall group) and those who did not (non-fall group). To assess the impact of various factors on fall occurrence, we excluded the cases of recurrent falls and focused our analysis solely on the first fall. Subsequently, a random-sampling approach was employed to balance the representations of both patient groups. The final stage of cohort refinement involved excluding the patients based on the completeness of the RTLS data. Patients were excluded if their RTLS records were unavailable throughout their hospitalization (n=167) or the RTLS data of fall patients were not recorded prior to the fall event (n=4). After establishing this refined cohort, we applied an 8:2 random split to separate the data into training and testing datasets (Fig. 2). This study was approved by the Institutional Review Board of Yonsei University Severance Hospital (No. 9-2021-0037).

### 4.2 Features engineering using clinical information extracted from EMR

We collected and selected the initial EMR data of all admitted patients, including the standard set of measurements performed during admission considering the anthropometric variables, such as height, weight, systolic blood pressure (SBP), diastolic blood pressure (DBP), and biochemical parameters including glucose, creatinine, alkaline phosphatase (ALP), alanine aminotransferase (ALT), aspartate aminotransferase (AST), bilirubin, blood urea nitrogen (BUN), calcium and lipid profiles, and complete blood count (CBC). The initial value was selected for model development in the case of repeated measurements. Hospitalization variables, such as the duration of admission and presence of ICU care, were also determined. For non-fall patients, the study period was from admission to discharge, whereas for fall patients, it was from admission to the day of the fall. Additionally, we considered the dosage of two types of drugs—peridol and sedatives—owing to their influence on the movement of patients, resulting in 27 attributes. The department codes were categorized into four types based on the medical department characteristics, as outlined in Supplementary Table 3. Descriptions and abbreviations of EMR features are provided in Supplementary Table 4.

### 4.3 RTLS-assisted feature engineering

Variables from the RTLS records were formulated to measure the dynamics of daily patient movements, as shown in Fig. 6. Fig. 6a illustrates an example of the RTLS dataset for a hypothetical patient, where *L* and *T* represent the recorded location of the patient and time point, respectively. This dataset serves as the foundation for subsequent feature engineering steps. In all equations, *k* denotes the k^th^ arbitrary row for each point of the same patient and *n* denotes the n^th^ arbitrary day for each patient.

**Fig. 6.**
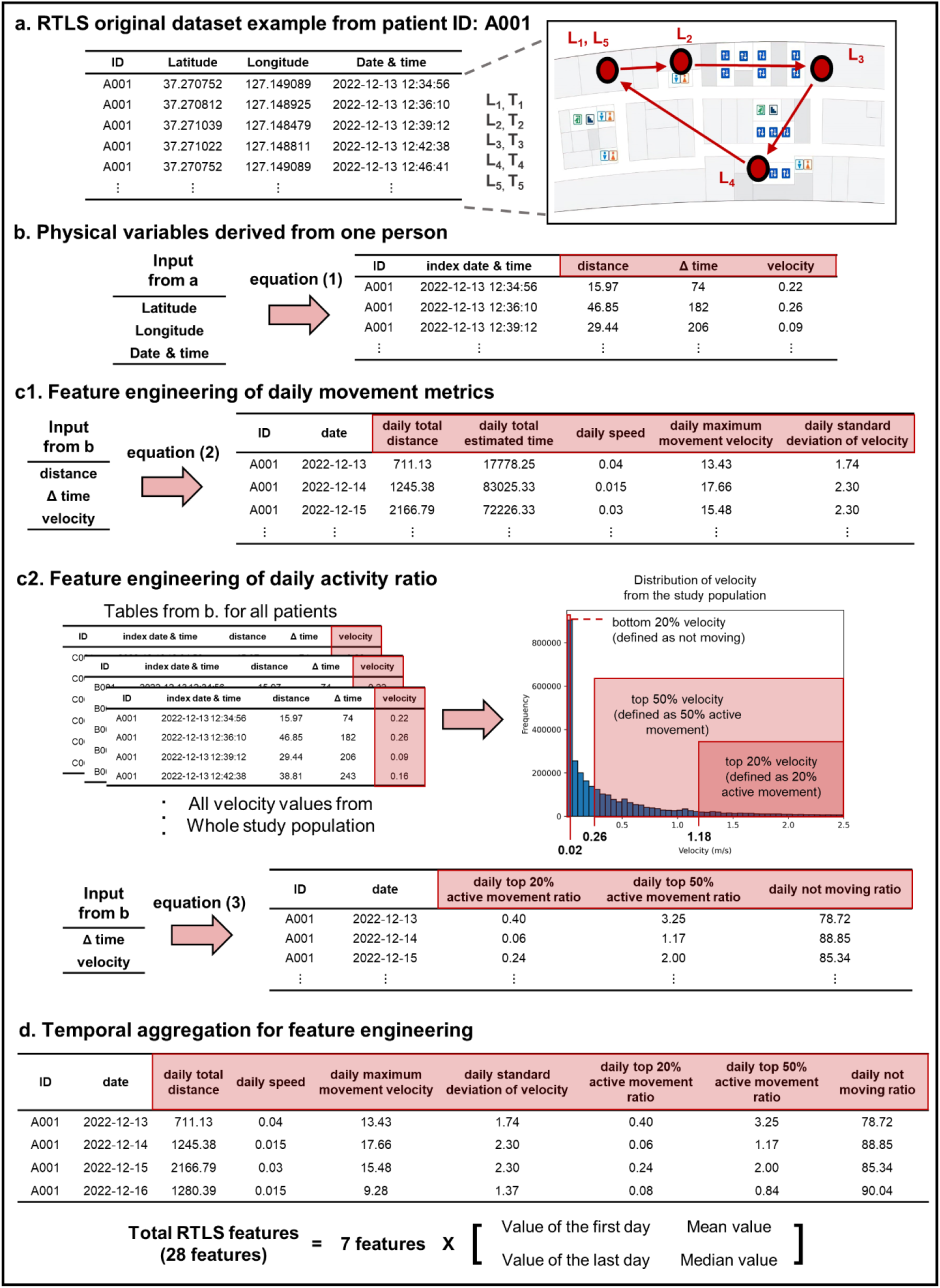
Overall flow of RTLS features engineering. a, Original RTLS dataset from patient ID A001. Example of a virtual patient’s movements across five locations within the facility (L1 to L5) tracked by the RTLS dataset following the sequence indicated by arrows. b, Physical variables derived from an individual. Calculation process for distance, Δ time, and velocity using the latitude, longitude, date, and time via Eq. 1 (1.1–1.3). c, Feature Engineering. c1: Feature engineering of daily movement metrics using distance, Δ time, and velocity to engineer daily total distance, daily total estimated time, daily speed, daily maximum movement velocity, and daily standard deviation of velocity using Eq. 2 (2.1–2.5), respectively. c2: Feature engineering of the daily activity ratio. Engineering the daily top 20% active movement ratio, daily top 50% active movement ratio, and daily not moving ratio using Δ time and velocity via Eq. 3 (3.1–3.3). Temporal aggregation for feature engineering. Aggregation of seven core features into 28 RTLS features by calculating the first and last day values, along with the mean and median values for each feature.

The differences between the consecutive rows in the dataset were initially calculated. The earlier date and time between two consecutive rows, referred to as the index date and time, respectively, served as the reference points. Here, Lat_k_ and Lng_k_ denote the latitude and longitude at location L_k_, respectively. Consequently, the variation in distance was redefined using the haversine formula, considering the Earth’s radius (r) as 6371 km (1.1). This formula accounts for the curvature of the Earth and provides a more accurate measurement of the distance between two points^29^. The change in time was defined as Δ time (1.2). The ratio of these two values is defined as velocity, which represents a vector incorporating the magnitude and direction of movement (1.3). The results presented in Fig. 6b were derived using these computations. For calculation, the units of distance and time were meters and seconds, respectively.

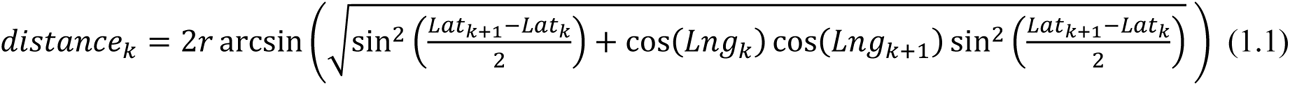

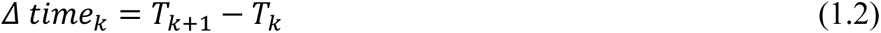

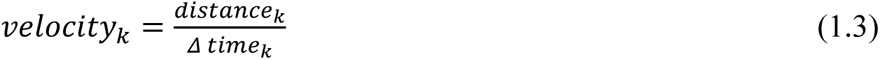

Subsequently, the basic metrics were aggregated to create features. The sums of daily distance and Δ time were defined as the daily total distance (2.1) and daily total estimated time (2.2), respectively. The ratio of these two sums was defined as the daily speed (2.3). Additionally, the daily maximum velocity and standard deviation were defined as the daily maximum movement velocity (2.4) and daily standard deviation of the velocity (2.5), respectively; the latter was used to examine fluctuations in patient movements^30^. These calculations yielded the results presented in Fig. 6c1. After excluding the total daily estimated time, the following four daily movement metrics were formulated:

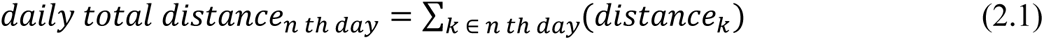

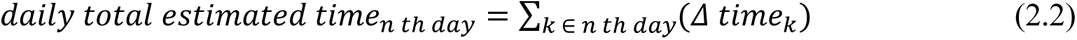

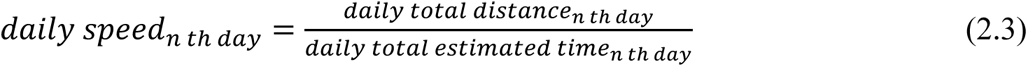

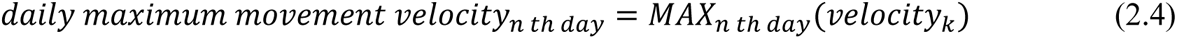

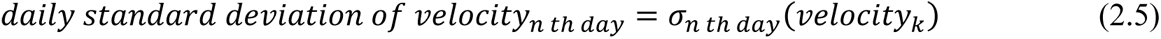

From the velocity data of the entire study population, we analyzed the distribution of all velocities and identified the following three significant thresholds: the top 20% of velocities at 1.18 m/s corresponding to the normal walking speed of adults^31^, median velocity at 0.26 m/s, and lower 20% of velocities at 0.02 m/s representing minimal movement. We calculated the duration for which each patient moved at a speed above these thresholds. Accordingly, the daily top 20% and 50% active movement ratios were defined as the duration for which each patient moved at a speed above 1.18 m/s (3.1) and 0.26 m/s (3.2), respectively. The daily not moving ratio (3.3) was defined as the duration for which each patient moved at a speed below 0.02 m/s, indicating minimal or no movement. The calculation results are shown in Fig. 6c2. The following three daily activity ratios were engineered based on the velocity thresholds:

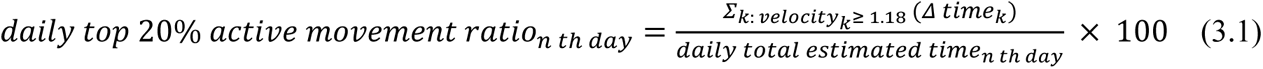

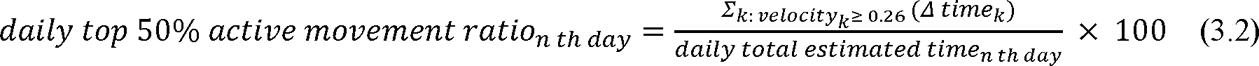

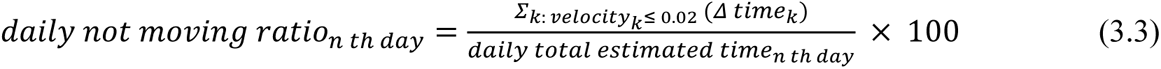

Finally, these metrics were compiled across multiple days during the stay of each patient (Fig. 6d). We included values from the first and last days, as well as the averages and medians of each feature over the duration to demonstrate the dynamics of patient movement. This resulted in a comprehensive set of 28 RTLS-based features for each patient. Further descriptions of each RTLS feature are summarized in Supplementary Table 5.

### 4.4 Data imputation

The process of enhancing the accuracy of in-hospital fall prediction commenced by addressing the issue of missing data, which is a common problem in data analysis that can lead to erroneous estimations. Detailed counts of missing values in the EMR data are presented in Supplementary Table 4. The KNN algorithm, known as the KNN imputation technique, was employed as the initial step to effectively impute missing values^32^.

### 4.5 Development and optimization of the prediction model

The modeling was performed using the XGBoost algorithm. To further refine the selected model, we incorporated 10-fold cross-validation (CV) and GridSearch for hyperparameter optimization. The Youden index was derived from an optimal model to assess the diagnostic ability. Using 27 clinical and 28 RTLS features, we developed the following three distinct models: the first using only clinical features (clinical model), the second using only RTLS features (RTLS model), and the third combining both the feature sets (clinical + RTLS model).

### 4.6 Performance assessment

To quantify and compare the performances of the three models on the test set, we employed bootstrapping with 1000 iterations to assess the performance at each iteration using various metrics. The performances were compared using the AUROC, AUPRC, and Brier scores. Subsequently, a detailed comparison was performed using the Youden index, which encompasses the accuracy, PPV, sensitivity, specificity, and F1 score.

Additionally, we employed DCA to compare the three models and determine clinical utility^33^. The net benefit refers to the balance between the true- and false-positive predictions for assessing the model’s predictive accuracy. Furthermore, SHAP values were used to identify the relative importance of the variables influencing the three models^34^.

### 4.7 Statistical analysis

To discern the statistical variations in patient characteristics, the data were first assessed using the Shapiro–Wilk test for normality. For binary attributes, the chi-square test was used to determine the statistical significance. For continuous variables, a suitable test was determined based on data distribution. The t-test and Mann–Whitney U test were applied when the data adhered to a normal and non-normal distribution, respectively. The Kruskal–Wallis test performed a nonparametric comparison of the distributions of outcome values across the three models. Subsequently, a post-hoc analysis was performed using the Bonferroni adjustment in conjunction with Dunn’s test to compare the significance across specific subgroups. Different levels of statistical significance are denoted in the plots as follows: an asterisk (*) for p-values less than 0.05, double asterisks (**) for p-values < 0.01, and triple asterisks (***) for p-values < 0.001.

## Author Contributions

DY and KMK conceptualized and specified the aims of the study. KMK and SK extracted medical records and labelled fall incidents. CMP administrated and extracted RTLS data. DWK and JS contributed to the study design, data preprocessing, hyperparameter tuning, development of the machine learning models, and writing the manuscript. CH and YK advised the study design and statistical analysis. DY and KMK supervised the project and reviewed and edited the manuscript. All authors have read and approved the final manuscript.

## Acknowledgements

This research was supported by a grant of the Korea Health Technology R&D Project through the Korea Health Industry Development Institute (KHIDI), funded by the Ministry of Health and Welfare, Republic of Korea (grant number: HI22C0452). And this work was supported by the National Research Foundation of Korea (NRF) grant funded by the Korean government (MSIT) (RS-2023-00253897) and grand funded by the Ministry of Education (2021R1I1A1A01060333).

## Data availability

The datasets used and/or analyzed in the current study are available from the corresponding author upon reasonable request.

## Code availability

The underlying code for this study is not publicly available but may be made available to qualified researchers upon reasonable request from the corresponding author.

## References

1. Florence, C. S. et al. Medical Costs of Fatal and Nonfatal Falls in Older Adults. J. Am. Geriatr. Soc. 66, 693–698 (2018).

2. Schwendimann, R., Bühler, H., De Geest, S. & Milisen, K. Falls and consequent injuries in hospitalized patients: effects of an interdisciplinary falls prevention program. BMC Health Serv. Res. 6, 69 (2006).

3. Fischer, I. D. et al. Patterns and predictors of inpatient falls and fall-related injuries in a large academic hospital. Infect. Control Hosp. Epidemiol. 26, 822–827 (2005).

4. Goodwin, M. B. & Westbrook, J. I. An analysis of patient accidents in hospital. Aust. Clin. Rev. 13, 141–149 (1993).

5. Morse, J. M. & Morse, R. M. Calculating fall rates: methodological concerns. QRB Qual. Rev. Bull. 14, 369–371 (1988).

6. Kim, M.-S., Jung, H.-M., Lee, H.-Y. & Kim, J. Risk Factors for Fall-Related Serious Injury among Korean Adults: A Cross-Sectional Retrospective Analysis. Int. J. Environ. Res. Public Health 18, (2021).

7. Matarese, M., Ivziku, D., Bartolozzi, F., Piredda, M. & De Marinis, M. G. Systematic review of fall risk screening tools for older patients in acute hospitals. J. Adv. Nurs. 71, 1198–1209 (2015).

8. Papaioannou, A. et al. Prediction of falls using a risk assessment tool in the acute care setting. BMC Med. 2, 1 (2004).

9. Barry, E., Galvin, R., Keogh, C., Horgan, F. & Fahey, T. Is the Timed Up and Go test a useful predictor of risk of falls in community dwelling older adults: a systematic review and meta-analysis. BMC Geriatr. 14, 14 (2014).

10. Presta, V. et al. Receiver Operating Characteristic Analysis of Posture and Gait Parameters to Prevent Frailty Condition and Fall Risk in the Elderly. NATO Adv. Sci. Inst. Ser. E Appl. Sci. 13, 3387 (2023).

11. Fazio, S. et al. How much do hospitalized adults move? A systematic review and meta-analysis. Appl. Nurs. Res. 51, 151189 (2020).

12. Growdon, M. E., Shorr, R. I. & Inouye, S. K. The Tension Between Promoting Mobility and Preventing Falls in the Hospital. JAMA Intern. Med. 177, 759–760 (2017).

13. Kyrdalen, I. L., Thingstad, P., Sandvik, L. & Ormstad, H. Associations between gait speed and well-known fall risk factors among community-dwelling older adults. Physiother. Res. Int. 24, e1743 (2019).

14. Thibaud, M. et al. Impact of physical activity and sedentary behaviour on fall risks in older people: a systematic review and meta-analysis of observational studies. Eur. Rev. Aging Phys. Act. 9, 5– 15 (2012).

15. Van Ancum, J. M. et al. Change in muscle strength and muscle mass in older hospitalized patients: A systematic review and meta-analysis. Exp. Gerontol. 92, 34–41 (2017).

16. Duan-Porter, W. et al. Hospitalization-Associated Change in Gait Speed and Risk of Functional Limitations for Older Adults. J. Gerontol. A Biol. Sci. Med. Sci. 74, 1657–1663 (2019).

17. Kim, M. H., Ryu, U. H., Heo, S.-J., Kim, Y. C. & Park, Y. S. The Potential Role of an Adjunctive Real-Time Locating System in Preventing Secondary Transmission of SARS-CoV-2 in a Hospital Environment: Retrospective Case-Control Study. J. Med. Internet Res. 24, e41395 (2022).

18. Yokota, S., Endo, M. & Ohe, K. Establishing a Classification System for High Fall-Risk Among Inpatients Using Support Vector Machines. Comput. Inform. Nurs. 35, 408–416 (2017).

19. Strini, V., Schiavolin, R. & Prendin, A. Fall Risk Assessment Scales: A Systematic Literature Review. Nurs Rep 11, 430–443 (2021).

20. Adam, C. E. et al. Change in gait speed and fall risk among community-dwelling older adults with and without mild cognitive impairment: a retrospective cohort analysis. BMC Geriatr. 23, 328 (2023).

21. Kulkarni, S. & Nagarkar, A. Basic gait pattern and impact of fall risk factors on gait among older adults in India. Gait Posture 88, 16–21 (2021).

22. Liang, C.-K. et al. Gait speed and risk assessment for falls among men aged 80 years and older: A prospective cohort study in Taiwan. Eur. Geriatr. Med. 5, 298–302 (2014).

23. Park, C., Atique, M. M. U., Mishra, R. & Najafi, B. Association between Fall History and Gait, Balance, Physical Activity, Depression, Fear of Falling, and Motor Capacity: A 6-Month Follow-Up Study. Int. J. Environ. Res. Public Health 19, (2022).

24. Michalcova, J., Vasut, K., Airaksinen, M. & Bielakova, K. Inclusion of medication-related fall risk in fall risk assessment tool in geriatric care units. BMC Geriatr. 20, 454 (2020).

25. Osman, A., Kamkar, N., Speechley, M., Ali, S. & Montero-Odasso, M. Fall risk-increasing drugs and gait performance in community-dwelling older adults: A systematic review. Ageing Res. Rev. 77, 101599 (2022).

26. Kim, K. M. et al. Red Cell Distribution Width Is a Risk Factor for Hip Fracture in Elderly Men Without Anemia. J. Bone Miner. Res. 35, 869–874 (2020).

27. Kim, K. M., Nerlekar, R., Tranah, G. J., Browner, W. S. & Cummings, S. R. Higher red cell distribution width and poorer hospitalization-related outcomes in elderly patients. J. Am. Geriatr. Soc. 70, 2354–2362 (2022).

28. Kim, K. M. et al. Association Between Variation in Red Cell Size and Multiple Aging-Related Outcomes. J. Gerontol. A Biol. Sci. Med. Sci. 76, 1288–1294 (2021).

29. Gao, J. et al. Evidence-driven spatiotemporal COVID-19 hospitalization prediction with Ising dynamics. Nat. Commun. 14, 1–14 (2023).

30. On the Use of Unsupervised Techniques for Fraud Detection in VoIP Networks. in Emerging Trends in ICT Security 359–373 (Morgan Kaufmann, 2014).

31. Sikandar, T. et al. Using a Deep Learning Method and Data from Two-Dimensional (2D) Marker-Less Video-Based Images for Walking Speed Classification. Sensors 21, (2021).

32. Beretta, L. & Santaniello, A. Nearest neighbor imputation algorithms: a critical evaluation. BMC Med. Inform. Decis. Mak. 16 **Suppl 3**, 74 (2016).

33. Clift, A. K. et al. Predicting 10-year breast cancer mortality risk in the general female population in England: a model development and validation study. The Lancet Digital Health 5, e571–e581 (2023).

34. Lundberg, S. M. et al. From Local Explanations to Global Understanding with Explainable AI for Trees. Nat Mach Intell 2, 56–67 (2020).

