## Supplementary table and figure for "Integrating Real-Time Location Systems with Electronic Medical Records: A Machine Learning Approach for In-Hospital Fall Risk Prediction"

**Supplementary materials**

**Table of contents**

***Supplementary Figures***

*Supplementary Fig. 1. Additional performance evaluation using the interval plots of the three models.*

*Supplementary Fig. 2. Feature importance of clinical and RTLS models using SHAP values.*

*Supplementary Fig. 3. RTLS sensors of the Yongin Severance hospital.*

***Supplementary Tables***

*Supplementary Table 1. Baseline characteristics (RTLS features) of patients.*

*Supplementary Table 2. Additional comparative performance metrics of the three models.*

*Supplementary Table 3. Department code classification.*

*Supplementary Table 4. Descriptions and counts of the missing clinical feature values.*

*Supplementary Table 5. Descriptions of RTLS features.*

**Supplementary Fig. 1. Additional performance evaluation using the interval plots of the three models.**

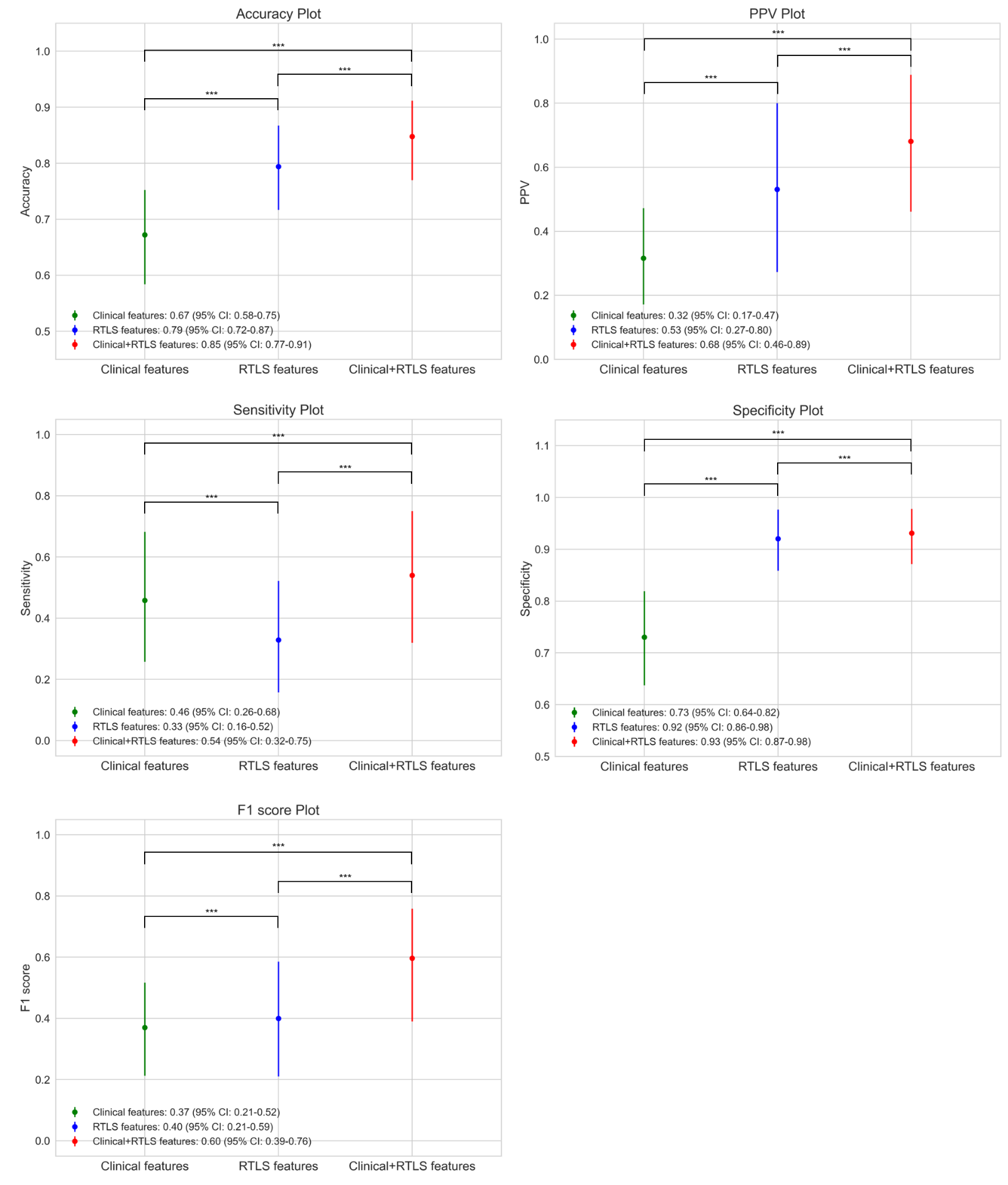

Performance comparison of the three models using accuracy, PPV, sensitivity, specificity, and F1 score based on 1000 bootstrap resamplings. The results are presented as 95% confidence intervals. Statistical significance is assessed using the Kruskal–Wallis test followed by Dunn's post-hoc test, with significance levels indicated on the graph. The clinical, RTLS, and clinical + RTLS models are denoted in green, blue, and red, respectively. PPV: positive predictive value; CI: confidence interval.

**Supplementary Fig. 2. Feature importance of clinical and RTLS models using SHAP values.**

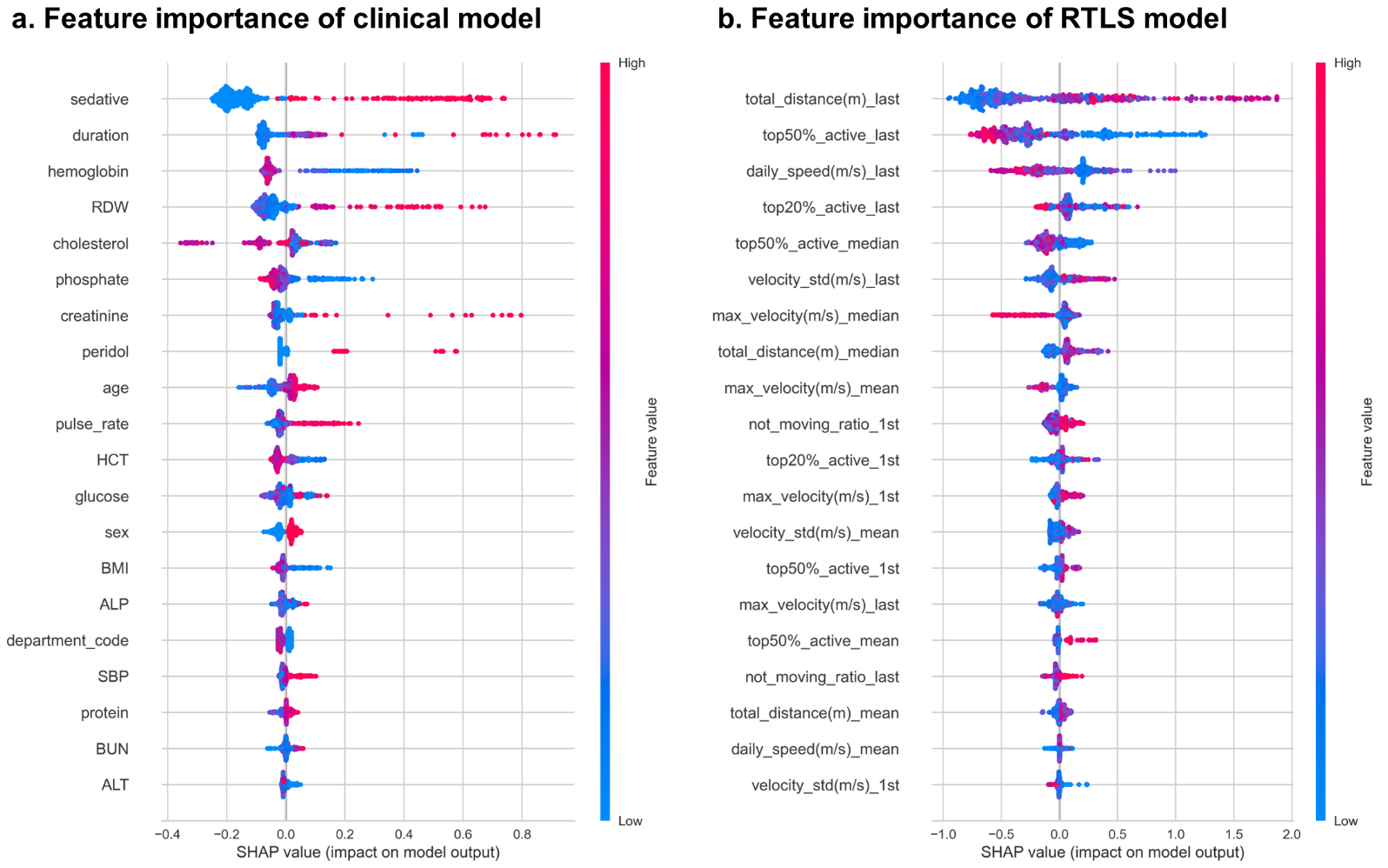

Impact of the top 20 features on model prediction ranked based on SHAP values. The y-axis displays the features in the order of decreasing impact, whereas the x-axis quantifies their SHAP values, with the color intensity ranging from blue for lower values to red for higher values. Specific descriptions of the clinical and RTLS features are listed in Supplementary Tables 4 and 5, respectively. Top 20 important features of the a, clinical model and b, RTLS model determined via SHAP analysis. RTLS: real-time location system; SHAP value: Shapley additive explanation value.

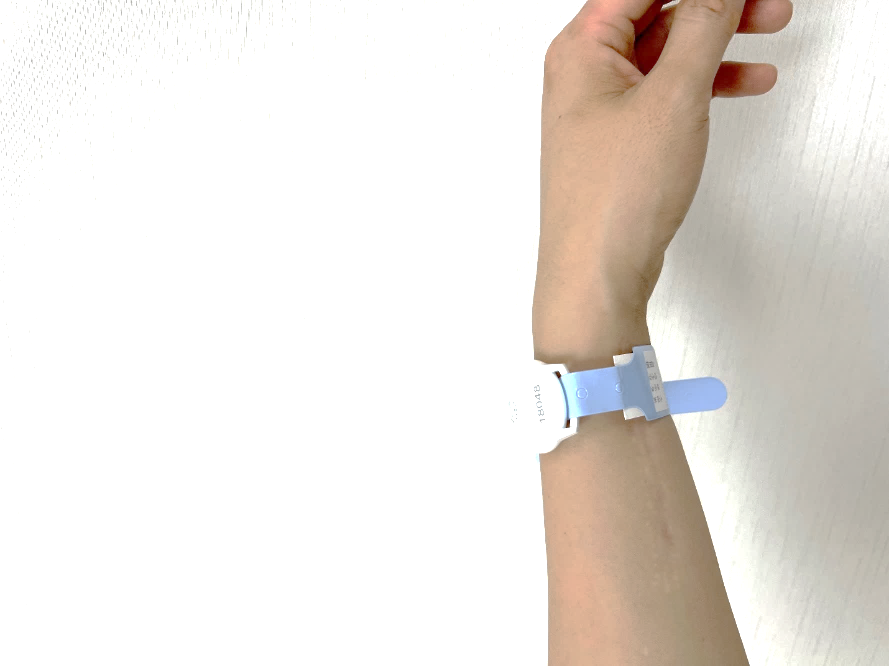
**Supplementary Fig. 3.** **RTLS sensors of the Yongin Severance hospital.**

**a. RTLS-equipped wristbands of patients.**

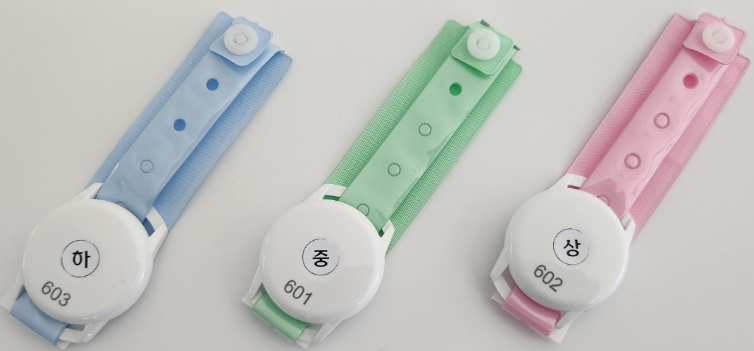

**b. On-model display of RTLS-equipped wristband.**

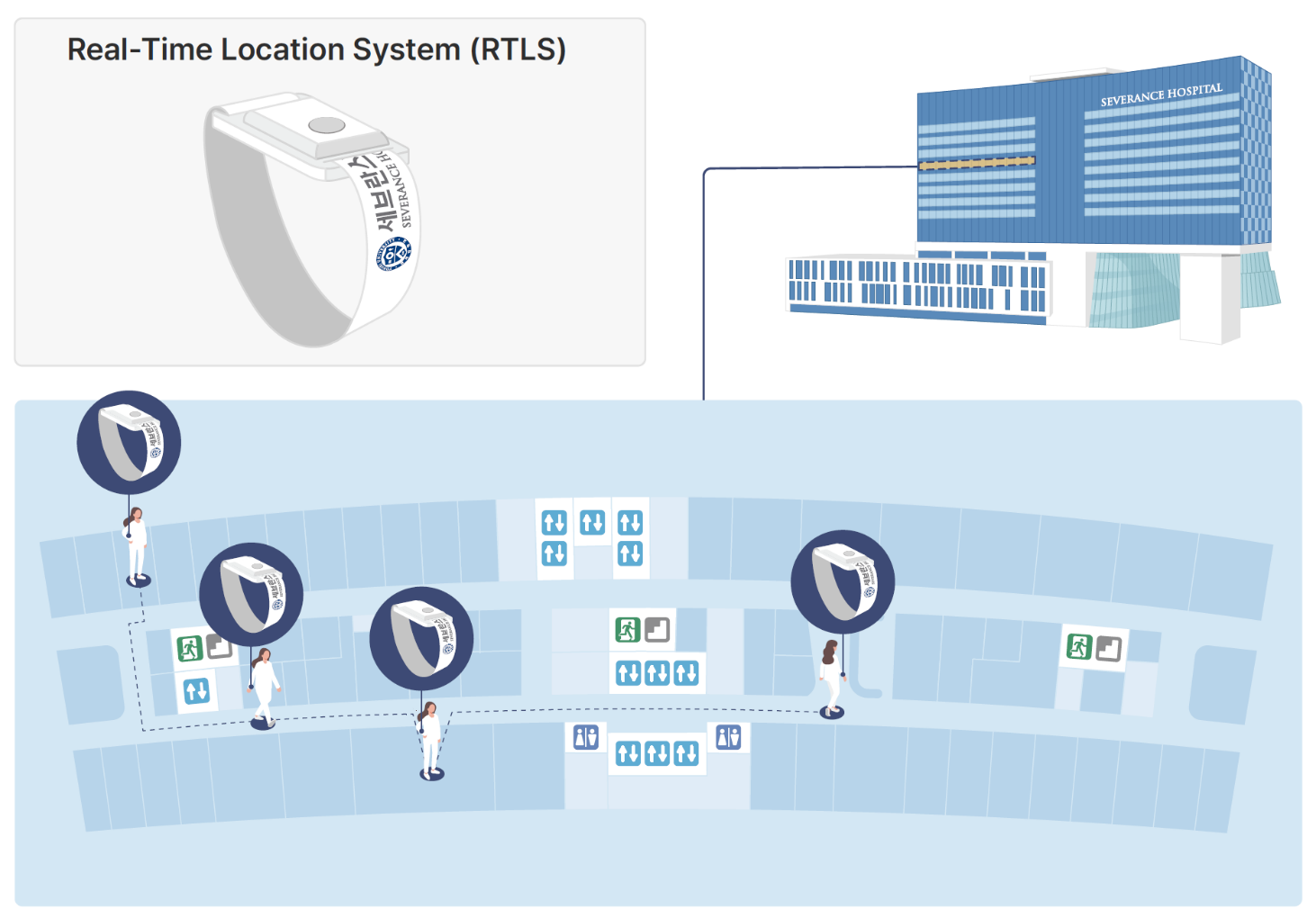

**c. Diagrammatic illustration of RTLS sensor measurement in Yongin Severance hospital.**

a, RTLS-equipped wristbands of patients serving as identification bracelets with embedded RTLS sensors. The wristbands are color-coded in blue, green, and red to represent patient risk levels, with each color indicating a different level of severity. b, Photograph of a patient wearing an RTLS-equipped wristband. c, Operational schematic of RTLS sensor tracking at Yongin Severance Hospital. The illustration above depicts a simplified blueprint of Yongin Severance Hospital. Below, the image illustrates the tracking of patient movement within a single floor, as recognized by RTLS wristband sensors. The RTLS technology identifies and records patient movement as they undergo medical examinations, receive treatments, or take rest periods, providing a real-time overview of patient activity on the premises. RTLS: Real-time location system.

**Supplementary Table 1.** **Baseline characteristics (RTLS features) of patients.**

|  | **Overall (n=561)** | **Fall (n=118)** | **No-fall (n=443)** | **P-value** |
| --- | --- | --- | --- | --- |
| **Daily total distance (m)** |  |  |  |  |
| **First day** | 1719.9 [700.7,3405.2] | 1773.0 [778.0,3261.9] | 1718.8 [685.8,3434.6] | 0.753 |
| **Last day** | 3710.8 [2049.5,5685.4] | 4509.5 [2351.7,6675.1] | 3640.8 [1990.3,5427.6] | 0.055 |
| **Average over LOS** | 4610.1 [2821.7,6530.0] | 4410.2 [2586.5,5979.2] | 4683.6 [2832.7,6640.5] | 0.225 |
| **Median over LOS** | 4173.3 [2408.9,6166.0] | 4055.2 [2196.2,6077.6] | 4228.0 [2484.1,6221.5] | 0.363 |
| **Daily speed (m/s)** |  |  |  |  |
| **First day** | 0.08 [0.04,0.13] | 0.08 [0.04,0.13] | 0.08 [0.04,0.14] | 0.819 |
| **Last day** | 0.07 [0.04,0.10] | 0.06 [0.03,0.09] | 0.07 [0.04,0.10] | 0.008 |
| **Average over LOS** | 0.07 [0.05,0.11] | 0.06 [0.04,0.09] | 0.08 [0.05,0.11] | 0.023 |
| **Median over LOS** | 0.07 [0.04,0.10] | 0.05 [0.03,0.09] | 0.07 [0.04,0.10] | 0.019 |
| **Daily max velocity (m/s)** |  |  |  |  |
| **First day** | 14.1 [8.2,23.5] | 15.0 [8.3,25.3] | 14.0 [8.2,21.9] | 0.582 |
| **Last day** | 18.0 [11.0,28.0] | 19.0 [10.9,27.8] | 17.5 [11.1,28.0] | 0.751 |
| **Average over LOS** | 20.3 [14.2,31.2] | 19.9 [14.5,29.3] | 20.4 [14.2,32.3] | 0.441 |
| **Median over LOS** | 17.9 [12.6,26.1] | 16.7 [12.5,24.2] | 18.1 [12.6,26.8] | 0.268 |
| **Daily SD of velocity** |  |  |  |  |
| **First day** | 2.7 (4.9) | 2.5 (2.5) | 2.7 (5.3) | 0.512 |
| **Last day** | 2.0 [1.4,2.8] | 2.1 [1.4,3.1] | 2.0 [1.4,2.6] | 0.2 |
| **Average over LOS** | 2.2 [1.7,3.2] | 2.3 [1.6,3.2] | 2.2 [1.7,3.2] | 0.854 |
| **Median over LOS** | 2.0 [1.6,2.8] | 2.0 [1.5,2.8] | 2.0 [1.6,2.9] | 0.676 |
| **Not moving ratio** |  |  |  |  |
| **First day** | 63.1 [40.0,80.7] | 63.0 [43.1,83.9] | 63.2 [38.9,80.3] | 0.573 |
| **Last day** | 72.1 [54.1,84.7] | 76.8 [55.9,89.0] | 71.2 [53.4,83.8] | 0.085 |
| **Average over LOS** | 70.8 [56.2,81.2] | 74.3 [56.1,82.8] | 69.7 [56.4,80.0] | 0.145 |
| **Median over LOS** | 72.2 [56.1,84.0] | 75.8 [55.6,86.5] | 71.2 [56.2,83.2] | 0.14 |
| **Daily top 50% active movement ratio** |  |  |  |  |
| **First day** | 5.8 [2.5,9.8] | 5.6 [2.5,9.9] | 5.8 [2.6,9.7] | 0.874 |
| **Last day** | 4.8 [2.4,7.5] | 3.4 [1.7,6.9] | 5.2 [2.7,7.8] | 0.001 |
| **Average over LOS** | 5.1 [3.0,7.6] | 4.6 [2.7,7.0] | 5.2 [3.3,7.7] | 0.081 |
| **Median over LOS** | 4.6 [2.5,7.2] | 3.5 [2.0,6.4] | 4.8 [2.7,7.3] | 0.012 |
| **Daily top 20% active movement ratio** |  |  |  |  |
| **First day** | 1.0 [0.3,2.2] | 1.1 [0.3,2.2] | 1.0 [0.3,2.2] | 0.967 |
| **Last day** | 0.9 [0.4,1.6] | 0.8 [0.4,1.3] | 0.9 [0.4,1.7] | 0.035 |
| **Average over LOS** | 1.0 [0.6,1.6] | 0.9 [0.5,1.4] | 1.1 [0.6,1.7] | 0.03 |
| **Median over LOS** | 0.9 [0.4,1.5] | 0.7 [0.4,1.2] | 1.0 [0.5,1.6] | 0.01 |

Data are presented as median (IQR) and mean (SD) for non-normally and normally distributed data, respectively. Units are specified next to each variable. The Shapiro–Wilk test was used to evaluate the normality of continuous variables. The t-test was employed for data satisfying the normality criteria, whereas the Mann–Whitney U test was applied for data not adhering to a normal distribution. LOS: length of stay; SD: standard deviation.

**Supplementary Table 2. Additional comparative performance metrics of the three models.**

| **Model** | **Youden index** | **Accuracy** | **PPV** | **Sensitivity** | **Specificity** | **F1 score** |
| --- | --- | --- | --- | --- | --- | --- |
| Clinical model | 0.298 | 0.672 (0.584-0.752) | 0.316 (0.171-0.472) | 0.458 (0.258-0.682) | 0.730 (0.637-0.819) | 0.370 (0.213-0.517) |
| RTLS model | 0.428 | 0.794 (0.717-0.867) | 0.581 (0.272-0.800) | 0.329 (0.158-0.522) | 0.921 (0.859-0.977) | 0.400 (0.211-0.585) |
| Clinical + RTLS model | 0.376 | 0.848 (0.770-0.912) | 0.681 (0.462-0.889) | 0.540 (0.320-0.750) | 0.931 (0.872-0.978) | 0.600 (0.390-0.759) |

The displayed values are the performance metrics for the clinical, RTLS, and clinical + RTLS models, with 95% confidence intervals (CIs) in parentheses. RTLS: real-time location system; PPV: positive predictive value.

**Supplementary Table 3. Department code classification.**

| **Department code** | **Department** |
| --- | --- |
| Medicine | Family Medicine / Infectious Disease / Internal Medicine / Endocrinology / Rheumatology / Gastroenterology / Nephrology / Cardiology / Physical Medicine and Rehabilitation / Psychiatry / Hematology-Oncology / Pulmonology and Allergy / Respiratory Allergy |
| Major surgery | Hepato-Biliary-Pancreatic Surgery / Thyroid and Endocrine Surgery / Colorectal Surgery / Obstetrics and Gynecology / Neurosurgery / General Surgery / Gastrointestinal Surgery / Breast Surgery / Otolaryngology / Orthopedic Surgery / Thoracic Surgery |
| Minor surgery | Oral and Maxillofacial Surgery / Urology / Plastic Surgery /  Ophthalmology / Otorhinolaryngology / Dermatology |
| Others | Health Promotion Center / Anesthesiology and Pain Medicine / Emergency Medicine / Nuclear Medicine |

Classification of patient admissions into four department codes based on their department of hospitalization.

**Supplementary Table 4.** **Descriptions and counts of the missing clinical feature values.**

| **Clinical variable** | **Feature name** | **Count of missing values** |
| --- | --- | --- |
| Sex | sex | 0 |
| Age | age | 0 |
| Duration period | duration | 0 |
| Department code | department_code | 0 |
| Intensive care unit (ICU) admission status | icu | 0 |
| Systolic blood pressure | SBP | 1 |
| Diastolic blood pressure | DBP | 1 |
| Pulse rate | pulse_rate | 1 |
| Body mass index (BMI) | BMI | 15 |
| Sedative intake status | sedative | 0 |
| Peridol intake status | peridol | 0 |
| Serum albumin level | albumin | 80 |
| Serum alkaline phosphatase (ALP) level | ALP | 85 |
| Serum alanine aminotransferase (ALT) level | ALT | 78 |
| Serum aspartate aminotransferase (AST) level | AST | 78 |
| Serum bilirubin level | bilirubin | 80 |
| Serum blood urea nitrogen (BUN) level | BUN | 73 |
| Serum calcium level | calcium | 82 |
| Serum total cholesterol level | cholesterol | 91 |
| Serum creatinine level | creatinine | 73 |
| Serum glucose level | glucose | 81 |
| Whole blood hematocrit level | HCT | 64 |
| Whole blood hemoglobin level | hemoglobin | 64 |
| Serum inorganic phosphate level | phosphate | 82 |
| Serum total protein level | protein | 80 |
| Whole blood red cell distribution width (RDW) | RDW | 64 |
| Serum uric acid level | uric_acid | 81 |

**Supplementary Table 5. Description of RTLS features.**

| **RTLS variable** | **Type** | **Feature name** | **Description** |
| --- | --- | --- | --- |
| Daily total distance (m) | First day | total_distance(m)_1st | Total distance moved (in meters) on the first day of RTLS records |
|  | Last day | total_distance(m)_last | Total distance moved (in meters) on the last day of RTLS records |
|  | Median | total_distance(m)_median | Median daily distance moved during hospitalization |
|  | Mean | total_distance(m)_mean | Average daily distance moved during hospitalization |
| Daily movement speed (m/s) | First day | daily_speed(m/s)_1st | Daily movement speed on the first day of RTLS records |
|  | Last day | daily_speed(m/s)_last | Daily movement speed on the last day of RTLS records |
|  | Median | daily_speed(m/s)_median | Median value of daily speed of movements during hospitalization |
|  | Mean | daily_speed(m/s)_mean | Mean value of daily speed of movements during hospitalization |
| Daily maximum movement velocity (m/s) | First day | max_velocity(m/s)_1st | Maximum velocity achieved on the first day of RTLS records |
|  | Last day | max_velocity(m/s)_last | Maximum velocity achieved on the last day of RTLS records |
|  | Median | max_velocity(m/s)_median | Median of daily maximum velocities recorded throughout hospitalization |
|  | Mean | max_velocity(m/s)_mean | Mean of daily maximum velocities recorded throughout hospitalization |
| Daily standard deviation  of velocity (m/s) | First day | velocity_std(m/s)_1st | Standard deviation of velocities on the first day of RTLS records |
|  | Last day | velocity_std(m/s)_last | Standard deviation of velocities on the last day of RTLS records |
|  | Median | velocity_std(m/s)_median | Median of daily velocity standard deviations throughout the hospitalization |
|  | Mean | velocity_std(m/s)_mean | Mean of daily velocity standard deviations throughout the hospitalization |
| Daily top 20% active movement ratio (%) | First day | top20%_active_1st | Ratio of time spent in the top 20% of active movements on the first day of RTLS records |
|  | Last day | top20%_active_last | Ratio of time spent in the top 20% of active movements on the last day of RTLS records |
|  | Median | top20%_active_median | Median of the daily ratio of time spent in the top 20% of active movements throughout the hospitalization |
|  | Mean | top20%_active_mean | Mean of the daily ratio of time spent in the top 20% of active movements throughout the hospitalization |
| Daily top 50% active movement ratio (%) | First day | top50%_active_1st | Ratio of time spent in the top 50% of active movements on the first day of RTLS records |
|  | Last day | top50%_active_last | Ratio of time spent in the top 50% of active movements on the last day of RTLS records |
|  | Median | top50%_active_median | Median of the daily ratio of time spent in the top 50% of active movements throughout the hospitalization |
|  | Mean | top50%_active_mean | Mean of the daily ratio of time spent in the top 50% of active movements throughout the hospitalization |
| Daily not moving ratio (%) | First day | not_moving_ratio_1st | Ratio of time without movement  on the first day of RTLS records |
|  | Last day | not_moving_ratio_last | Ratio of time without movement  on the last day of RTLS records |
|  | Median | not_moving_ratio_median | Median of the daily ratios of time  without movement throughout the hospitalization |
|  | Mean | not_moving_ratio_mean | Mean of the daily ratios of time  without movement throughout the hospitalization |

RTLS: real-time location system.
